# Simplified Co-extraction of total Nucleic Acids from Respiratory Samples for detection of *Mycobacterium tuberculosis* and SARS-CoV-2 optimized for compatibility across Diagnostic Platforms

**DOI:** 10.1101/2025.02.27.25322880

**Authors:** Nisha H. Modi, Owen R. S. Dunkley, Alexandra G. Bell, Emily Hennig, Aanchal Wats, Yujia Huang, Naranjargal Daivaa, Cameron Myhrvold, Yingda L. Xie, Padmapriya Banada

**Affiliations:** Department of Medicine, International Center for Public Health, Rutgers University, Newark, NJ, 07103; Department of Molecular Biology, Princeton University, Princeton, NJ, 08544; Department of Chemical and Biological Engineering, Princeton University, Princeton, NJ 08544; Omenn-Darling Bioengineering Institute, Princeton University, Princeton, NJ 08544; Department of Chemistry, Princeton University, Princeton, NJ 08544

## Abstract

Tuberculosis (TB) and COVID-19 are leading infectious diseases with high mortality, caused by *Mycobacterium tuberculosis* (*Mtb*) and *SARS-CoV-2 (SC2)*, respectively. Co-infection is common but is often undiagnosed as it is challenging to process both pathogens from a single sample. In this study, we present a simple and efficient method for co-extracting nucleic acids (NA) from these two distinct respiratory pathogens for downstream diagnostic testing. We evaluated three different nucleic acid amplification (NAA)-based platforms, LightCycler480 (LC480) qPCR, Qiacuity digital PCR (dPCR), and Cytation3 for CRISPR-Cas13a-based SHINE-TB/SC2 detection assays. Chelex-100 chelating resin-based boiling preparation method was optimized for *Mtb* NA extraction from saliva and sputum. Saliva showed compatibility with all three platforms, with sensitivity as low as 100 CFU/ml (or 2 genomic copies/µl). This method worked well for sputum using dPCR at 100% (21/21) positivity, though the CRISPR-based SHINE-TB assay showed more variability and sensitivity to sputum inhibitor carry-over, resulting in an 81% positive rate (17/21). Diluting sputum with TE buffer (1:1) improved the detection (2/4). Extraction efficiency of our method was 48%, 62.2%, 86.4% and 99.3% for concentrations 10^5^, 10^4^, 10^3^ and 10 CFU/ml, respectively. The dynamic range for *Mtb* spiked in pooled sputum showed 100% detection (N=8) at ≥10^3^ CFU/ml with all three methods. Dual-pathogen co-extraction and detection of *SC2* (10^5^ PFU/ml) and *Mtb* (10^5^ CFU/ml) in salivary sputum was successful using CRISPR-Cas13a assays. We have developed a rapid and efficient co-extraction method for multi-pathogen testing across diagnostic platforms and believe this is the first protocol optimized to co-extract *Mtb* and SARS-CoV-2 from a single sample.

## INTRODUCTION

Tuberculosis (TB) and COVID-19 are two of the deadliest infectious diseases worldwide, affecting millions of people each year. TB is caused by *Mycobacterium tuberculosis (Mtb),* an acid-fast bacterium while COVID-19 is caused by a coronavirus *SARS-CoV-2 (SC2)* (1). COVID-19 has led to >7 million deaths worldwide, and continues to be a major health challenge as new variants emerge (2). In 2023, an estimated 10.8 million people developed TB, with 8.2 million diagnosed and 1.25 million deaths reported (3, 4). TB and COVID-19 remain significant public health crises, especially in endemic countries where medical resources, treatment, and community interventions are scarce (5–7). The global efforts to eliminate TB were largely set back due to COVID-19 pandemic, which led to a rise in undiagnosed TB cases due to diversion of public health intervention programs away from TB towards COVID-19 (3, 8–11). This highlights the need for scalable multi-pathogen surveillance approaches implementable in community clinics for effective management and control of epidemics (12, 13) currently and in future.

COVID-19 and TB are respiratory diseases that primarily affect the lungs and can be diagnosed using respiratory samples (1, 14). Sputum is commonly used for diagnosing pulmonary TB while nasopharyngeal, nasal and saliva are often preferred for *SC2* (15–17). It is possible that a single sample type could be used for detection of both pathogens (12). Sputum can be difficult to collect from TB patients especially in vulnerable populations such as HIV positive, children, and the elderly. Often, multiple sample collections or invasive procedures are required to obtain a high-quality sputum sample, highlighting the need for alternative sample types for diagnostics. Various sample types including saliva, have been explored in recent years with promising results for pulmonary TB diagnosis (18, 19). Saliva has offered comparable sensitivity to sputum in lab developed tests (18). Similarly, sputum samples have showed equal or better sensitivity than oropharyngeal samples for COVID-19 testing (15), suggesting that either saliva or sputum could serve as single collection type for both TB and COVID-19 diagnosis.

Nucleic acid-based (NA) molecular diagnostic tests have significantly improved TB and COVID-19 diagnosis in terms of time, sensitivity, specificity and overall cost-effectiveness compared to traditional methods (14, 20–22). A variety of NA-based tests, including those developed by our team, have proven highly efficient for detecting TB and COVID-19 (23–30). However, there remains a gap in achieving optimal diagnostic yield, where the test accurately identifies all individuals with the infection (31). This gap could be due to the sample type requirements for certain tests or inefficient nucleic acid extraction limiting the test’s performance (32–35). To facilitate timely diagnosis in resource-limited settings, nucleic acid extraction from sample types containing mixed pathogens must be rapid, efficient, simple, and cost-effective.

In this study, we demonstrate the optimization and development of a simple total nucleic acid (DNA and RNA) extraction method by adapting the commonly used Chelex-100 resin and boiling technique. We have explored extraction of total nucleic acids from saliva and salivary sputum sample spiked with TB and SARS-CoV-2 and demonstrated compatibility across various tests and platforms. We propose a simple protocol that eliminates the need for specialized equipment, enabling streamlined multi-pathogen testing in community centers. To the best of our knowledge, no study has previously attempted to co-extract total nucleic acids from *Mtb* and *SC2* using the same sample type.

## METHODS

### Ethics statement

All discarded and de-identified patient samples were collected under the approval from the Rutgers Institutional review board protocol number Pro2020001138.

### Bacterial cultures

An attenuated strain of *M. tuberculosis* H37Rv (mc^2^6230, *Mtb*), and a vaccine strain *M. bovis* BCG (BCG) were used in all the experiments here unless indicated. Both were cultured in BD DIFCO™ 7H9 Middlebrook broth supplemented with 10% Middlebrook OADC growth supplement (BD) and 0.05% Tween 80 (Sigma Aldrich, St Louis, MO) as per the manufacturer’s recommendations (BD, Franklin Lakes, NJ). *M. tuberculosis* H37Rv (mc^2^6230, a kind gift from Dr. William Jacobs, Jr., Albert Einstein College of Medicine, Bronx, NY), has independent deletions in the *pan*C and *pan*D genes and requires media supplemented with 24 µg/ml of Calcium Pantheonate (Sigma Aldrich, St Louis, MO). Both strains were then grown to an optical density OD of 0.6-0.8 and sub-cultured twice and quantitated by plating on Middlebrook 7H10 agar plates (BD). The grown culture stock was mixed, aliquoted and stored at -80°C until further use. For spiking experiments, an aliquot of the quantified stock culture was thawed at 4°C, sonicated twice for 30s (Branson CPX1800-E Ultrasonic water bath, Danbury, CT) to ensure breakage of clumps and uniform distribution of cells. Ten-fold serial dilutions were made in 7H9 broth and spiked at required concentrations to the matrix (either pooled or individual sputum/saliva collected from non-TB suspects).

### Clinical Samples

Saliva and sputum were used as clinical matrix for this evaluation. Samples were collected from 39 participants at the University Hospital (UH) in Newark, NJ, USA between May 2022 and November 2023 <u>under an approved IRB protocol (Rutgers eIRB # Pro2020001138)</u>. Participants were diagnosed with non-TB conditions, including non-respiratory conditions (foot laceration, femur fracture, etc) or respiratory conditions (e.g. COPD, asthma, fluid overload due to congestive heart failure) who could produce expectorated sputum. Clinical characteristics, including gender, age, and underlying conditions, are shown in Supplementary Table 3. Samples were characterized based on physical properties, logged, aliquoted, and stored at -80°C until use. The use of the terminology salivary sputum refers to 1:1 diluted saliva in sputum. All individual sputum samples were tested by IS6110-dPCR to confirm that they were *Mtb*-negative before spiking with *Mtb*.

### Optimization of sample processing protocol

Sample processing buffer was designed using Chelex-100 resin based Instagene Matrix (Biorad) as the base reagent. We explored supplementing Chelex-100 with various detergents such as Nonidet P-40 (NP-40, Sigma), Tween20 (Sigma), Tergitol (Sigma), and TritonX (Sigma) at 1% concentration, for enhanced nucleic acid recovery. Negative saliva obtained from BioIVT, Westbury, NY (from 4 known, *Mtb*-negative participants), and was confirmed again at our lab using Xpert MTB/RIF ultra test. All samples were pooled at equal volumes and mixed via snap vortexing. Sputum collected from discarded patient samples from the microbiology lab at the University Hospital, Newark, NJ were pooled separately based on physical characteristics to evaluate the variability of the sputum sample type. The pools were verified negative by Xpert MTB/RIF Ultra test (Ultra, Cepheid, Sunnyvale, CA).

The initial protocol involved adding 200µl of the Chelex based buffer to 100µl of spiked (*Mtb* at ∼10^6^ CFU/ml) saliva/sputum in a screw-cap Eppendorf tube with an O-ring. The samples were vortexed for 30s at 3400 rpm, incubated for 30 min at 95°C, and then centrifuged for 2 min at 9400 xg. The supernatant containing the nucleic acid was carefully collected (∼100µl). All the extracted samples were analyzed via the qPCR IS6110 assay, dPCR IS6110 assay and the CRISPR-Cas13a IS6110/IS1081 assay.

### Assays Used for Validation

Primer and probe sequences for different assays used here are mentioned in Table S2, where IS6110 assay (23), and *acr* assay (36) were used as targets for *M. tuberculosis* detection and N1 assay (37) for SARS-CoV-2 .

The QIAcuity ProbePCR kit (Qiagen, Hilden, Germany) was used for QIAcuity Digital PCR System. The reaction components varied based on the nanoplate type, but the standardized kit-recommended reaction mix and cycling conditions were utilized. For 8.5K nanoplates (Qiagen, Hilden, Germany), the 12µl reaction mix was comprised of 3µl of PCR Master Mix (Qiagen, Hilden, Germany), 0.8uM of each primer, 0.4µM of probe, 5.8µl of water; and 2µl of DNA. For 26K nanoplates (Qiagen, Hilden, Germany), the 40µl reaction mix was comprised of 10µl of ProbePCR Master Mix, 0.8µM of each primer, 0.4µM of probe, 5.8µl of water; and 2µl of DNA. The dPCR cycling conditions were followed as per the manufacturer’s instructions (Qiagen), which included an initial heat activation at 95°C for 2 min, then 40 cycles of denaturation at 95°C for 15s, and annealing/extension at 60°C for 30s.

Real time PCR was performed in the LightCycler 480 system, in a 384 well microliter plate at a reaction volume of 12 µl (using the QIACuity probe PCR kit), comprised of 3µl of ProbePCR Master Mix (Qiagen, Hilden, Germany), 0.8µM of each primer, 0.4µM of acr/*IS6110* assay probe, 5.8µl of water; and 2µl of DNA. The PCR cycling conditions included an initial heat activation at 95°C for 2 min, then 40 cycles of denaturation at 95°C for 15s, and annealing at 60°C for 30 s. and extension at 72°C, 15 s. The fluorescence was recorded during the annealing step of the assay.

CRISPR SHINE-TB test was performed using the single-step Cas13a assay developed by our team (38) and CRISPR SHINEv.2 SARS-CoV-2 test (20). Briefly, SHINE-TB test master mix contained SHINE buffer (20 mM HEPES pH 8.0, 60 mM KCl, 3.5% PEG-8000, nuclease-free (nf) water), 45 nM *Lwa*Cas13a (Genscript, stored in 100 mM Tris HCl pH 7.5 and 1 mM DTT), 0.3 nM of each rNTP (New England Biolabs), 1 *U*/mL T7 RNAP (Biosearch Technologies), 62.5 nM FAM 6U quenched reporter (5’-/56-FAM/rUrUrUrUrUrU/3IABkFQ/-3’; IDT), 22.5 nM of each crRNA in a combination, 70 nM of the appropriate RPA primers in nf water and finally, 14 mM MgOAc. Assays were performed by mixing completed Cas13a master mix (90% by volume) with target (10% by volume). Reactions were loaded in technical triplicates into 384-well clear-bottom microplates (Greiner) for 15 µL reactions or 20 µL reactions in 384-well microplates (Corning). The plates were incubated at 37°C for up to 3 hours in an Agilent BioTek Cytation 5 microplate reader, with fluorescence readings programmed to record every 5 minutes.

### Extraction efficiency

*M. bovis* BCG was spiked to pooled sputum at different concentrations at 10^5^, 10^4^, 10^3^ and 10 CFU/ml and total nucleic acids were extracted following the protocol described in Fig. 2a. The dPCR was performed using IS6110 assay following the method explained earlier. Extraction efficiency was calculated using the formula Extraction Efficiency (%) = (DNA quantity recovered / Initial DNA quantity) x 100.

## RESULTS

### Optimization of Chelex-100 resin based sample processing for saliva and sputum

To evaluate the effect of different detergents commonly used to improve extraction yield from bacteria, the Chelex-100 resin (Instagene matrix) was supplemented with various detergents (Fig. 1) and tested on individual saliva and two different sputum samples (Fig. 1c) spiked with *Mtb* at 10^6^ CFU/ml. DNA extracted using boil preparation method described in the methods was evaluated with dPCR (Fig. 1a), and the CRISPR-based SHINE-TB test (Fig. 1b). The results showed no significant difference (P>0.05) between Chelex-100 with and without detergents for sputum pools A and B. However, for saliva, an improvement was observed with the addition of NP-40, Tergitol, and Triton-X-100 (Fig. 1a). With the CRISPR SHINE-TB assay, the yield of sputum B was no different compared to that of saliva in all conditions (P>0.05, Fig. 1b). Conversely for sputum A, the test failed across all Chelex +/- resin-based solutions (Fig. 1b), which could be due to the presence of inhibitors in sputum A, which was more darkly colored and mucoidal than sputum B (Fig. 1c). Considering the natural biological variation expected with sputum samples, further testing and optimization were necessary to efficiently detect *Mtb* from sputum in CRISPR-based tests.

**Fig 1.**
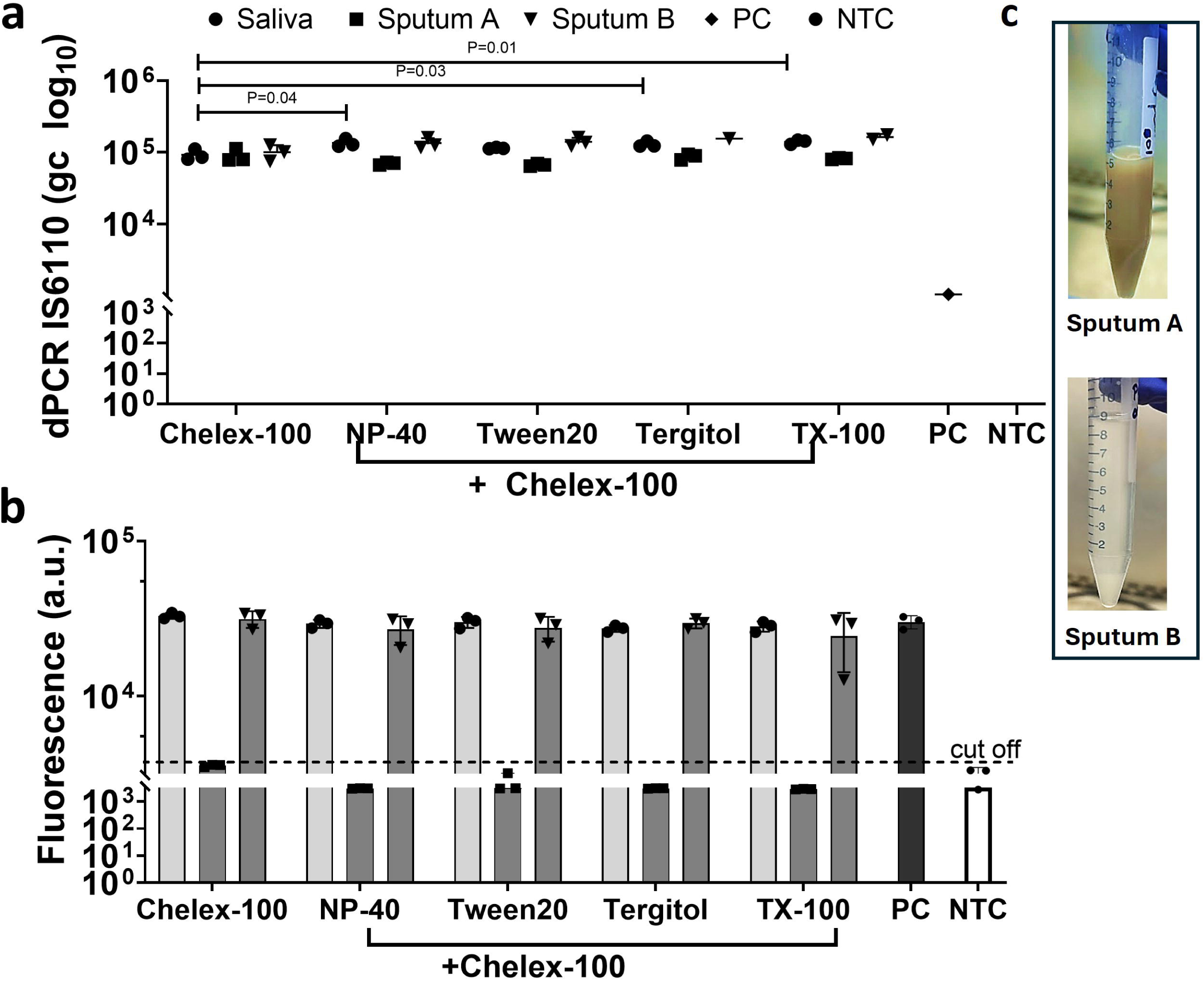
Optimization of Chelex-100 resin based sample processing. Chelex-100 was supplemented with various detergents, and total DNA was extracted by boil prep and the eluant was tested with a) digital PCR and b) SHINE-TB cas13a assay (fluorescence at 90 min (FAM, λEx. 490 nm/ λEm. 517nm)); c) Difference in the physical appearance of the two sputum samples sputum A and sputum B is shown. Sp. Sputum; PC-Positive control ATCC Mtb H37Ra at 10^3^ copies/reaction; NTC-No template control. Sputum samples A and B after pooling is shown. All conditions were compared against Chelex-100 control and P values to denote statistical significance is shown only for those samples with P<0.05, all other conditions have P>0.05.

### Saliva sample processing and detection in NAA-based methods

A confirmed negative saliva sample pool (BioIVT, NY) was spiked with *Mtb* H37Rv mc^2^6230 at logarithmic dilutions ranging from 10^6^ CFU to 100 CFU/ml. *Mtb* DNA was extracted from saliva using the optimized Chelex resin-based boil prep (CRB) protocol as described in Fig. 2a. The extracted DNA from different mycobacterial concentrations was tested across a variety of *Mtb* detection assays using different platforms, demonstrating compatibility across multiple methods. Real-time PCR targeting the *Mtb acr* gene was performed using the Roche LC480 system (Fig. 2b), while digital PCR (dPCR) on the Qiacuity platform (Fig. 2c) and the CRISPR-based SHINE-TB test (Fig. 2d and e) targeted the *Mtb IS6110* gene. Detection sensitivity varied between assays. The *acr* assay, which targets a single-copy gene, showed a sensitivity of 100% (9/9) at concentrations greater than 10L CFU/ml but dropped below 50% at 10³ CFU/ml (4/9) and 10² CFU/ml (5/9) of *Mtb* spiked in saliva.

**Fig 2.**
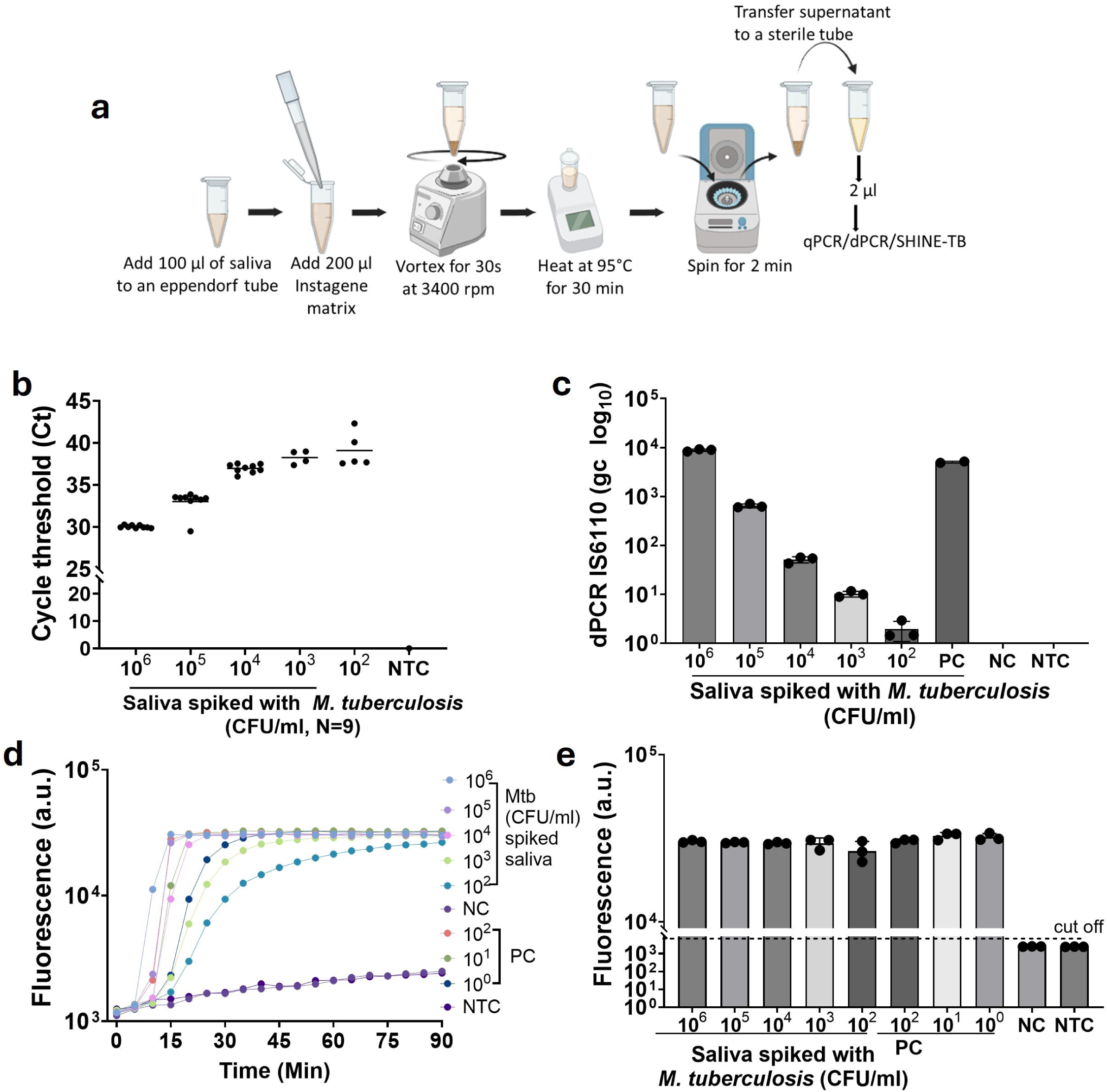
Saliva sample processing and eluant compatibility across various platforms. a) Optimized Chelex-100 resin based boil preparation (CRB) sample processing protocol for saliva; dynamic range for total DNA extracted using the protocol with saliva tested in b) Realtime PCR (*acr* gene) in Roche LC480 (N=9), c) Qiacuity dPCR (IS6110); d) and e) SHINE-TB cas13a assay (IS6110) in Cytation 3 platform showing d) real time curves e) end point fluorescence at 90 min (FAM λEx. 490 nm/ λEm. 517nm); Sp. Sputum; PC-Positive control ATCC *Mtb* H37Ra (used at 10^2^ copies/reaction in dPCR); NC-non spiked saliva extraction negative control; NTC-No template reaction control; a.u- arbitrary units.

However, the dPCR and SHINE-TB tests, both targeting the multi-copy *IS6110* gene, exhibited a 100% sensitivity at 100 CFU/ml (Fig. 2). These results indicate that *Mtb* DNA extracted from saliva using our optimized protocol is free of inhibitors and applicable across a broad range of platforms and *Mtb* detection assays.

### Sputum sample processing optimization for CRISPR-SHINE TB test

Sputum is a widely used and accepted sample type for TB testing and as shown in Fig. 1, we observed variability in detection with different sputum pools, primarily affecting SHINE-TB more than qPCR or dPCR. To assess this, we first evaluated 10 individual sputum samples collected from 10 different TB-unsuspected patients using the SHINE-TB test (Fig. 3a) using our CRB protocol mentioned in Fig. 2a. Remarkably, 5 out of 10 samples failed to detect *M. bovis* BCG at a concentration of 10L CFU/ml. We suspected two possible reasons for this failure: (1) insufficient mycobacterial lysis or (2) the presence of assay-specific inhibitors affecting Cas or RPA activity.

**Fig 3.**
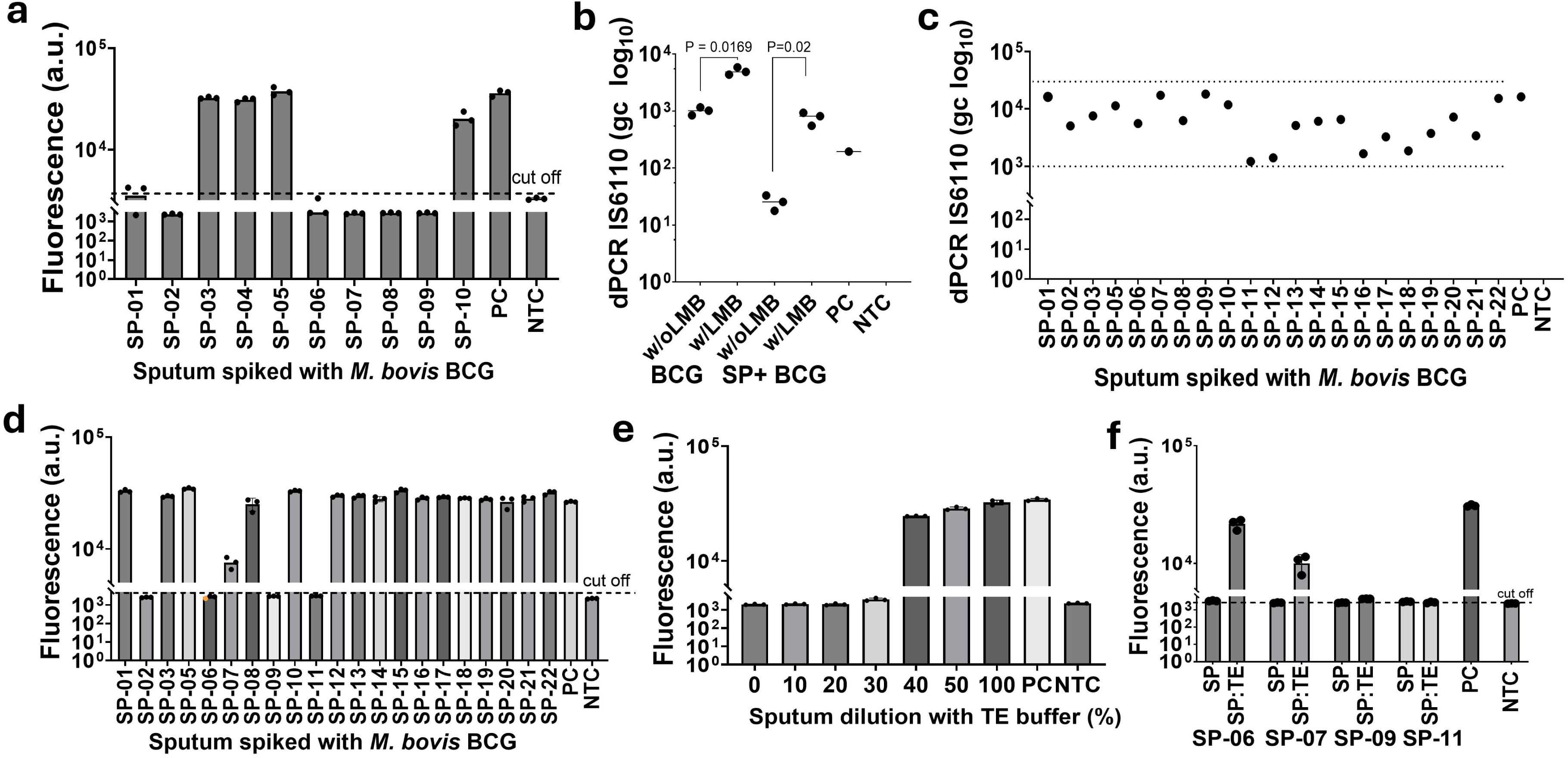
Sputum sample processing and reduction of sample inhibition in SHINE-TB assay. a) CRB sample processing method was used to extract total DNA with various negative patient sputum samples spiked with *M. bovis* BCG (10^5^ CFU/ml) and was tested with SHINE-TB assay; b) optimization of CRB method in dPCR supplementing with lysing matrix beads (LMB); c) evaluation of BCG spiked (10^5^ CFU/ml) individual patient sputum samples with optimized CRB+LMB method; d) Re-evaluation of SHINE-TB assay with BCG spiked (10^5^ CFU/ml) individual patient sputum samples with optimized CRB+LMB method. e) effect of dilution on sputum samples using Tris-EDTA buffer evaluated with SHINE-TB assay; f) evaluation of inhibitory samples using TE dilution method. a), d), e) and f) SHINE-TB test end point fluorescence units at 90 min (FAM λEx. 490 nm/ λEm. 517nm). Sp. Sputum; PC-BCG (10^5^ CFU/mL) spiked saliva extraction control; NTC-No template reaction control; a.u- arbitrary units.

To address the issue of insufficient lysis, we tested ready-to-use 2 ml tubes containing Lysing Matrix B beads (LMB, MP Biomedicals), which are 0.1 mm silica spheres designed to enhance *Mtb* lysis efficiency. *M. bovis* BCG was spiked at 10L CFU/ml into a pooled sputum sample composed of the five previously undetected samples (Sp-02, Sp-06, Sp-07, Sp-08, and Sp-09). DNA was then extracted using a modified CRB protocol (Fig. 2a), where LMB tubes replaced standard Eppendorf tubes. A 2 µl aliquot of the extracted DNA was tested using dPCR.

As shown in Fig. 3b and Fig. S1, the use of LMBs resulted in a 2-logLL increase in BCG copy number in spiked sputum compared to extractions without LMBs, demonstrating the positive effect of the beads on bacterial lysis and DNA yield. Further dPCR testing with 21 individual sputum samples spiked with BCG consistently showed higher detection rates when LMBs were used, with all samples (21/21) successfully detected. However, genomic copy numbers varied from 1,208 to 18,100, indicating some variability in extraction efficiency among samples.

When the same set of 21 samples was tested using the CRISPR SHINE-TB assay, 17 out of 21 samples (81%) were detected, while four samples (Sp-02, Sp-06, Sp-09, and Sp-11, Fig. 3d) remained undetected. Improper lysis or extraction was unlikely to be the cause of these failures, as dPCR confirmed that all undetected samples, except Sp-11, contained ≥10L genomic copies per reaction, well above the established limit of detection (LoD) for SHINE-TB (<100 CFU/ml, (38)). Sp-11 showed a lower copy number in dPCR (Fig. 3c), which may partially explain its failure to be detected.

Given these findings, we suspected the presence of inhibitors specific to RPA or Cas enzymes as the potential reason for assay failure. To test this hypothesis, we applied a simple dilution approach using TE buffer. We found that when the sample-to-TE buffer ratio exceeded 40% (i.e., 40% TE buffer and 60% sputum), SHINE-TB positivity was restored (Fig. 3e), supporting our hypothesis. Further experimentation is required to determine whether RPA- or Cas-specific inhibition is the primary factor. However, we were unable to conduct additional tests with the specific inhibitory sputum samples due to limited sample volume.

Nevertheless, we selected a 1:1 sputum-to-TE buffer dilution for further testing of inhibitory sputum samples from Fig. 3d. This dilution approach had somewhat positive impact, where 2 of the 4 samples tested positive (Fig. 3f). Further optimization and purification may be necessary to improve sputum testing with CRISPR-Cas-based assays.

Using this simple boil preparation method in combination with lysis beads and dilution (CRB, Fig. 4a), we established that the eluate from saliva and salivary sputum is compatible with a variety of platforms, including qPCR (LC480), dPCR (Qiacuity), and CRISPR-based SHINE-TB assays (Fig. 4).

**Fig 4.**
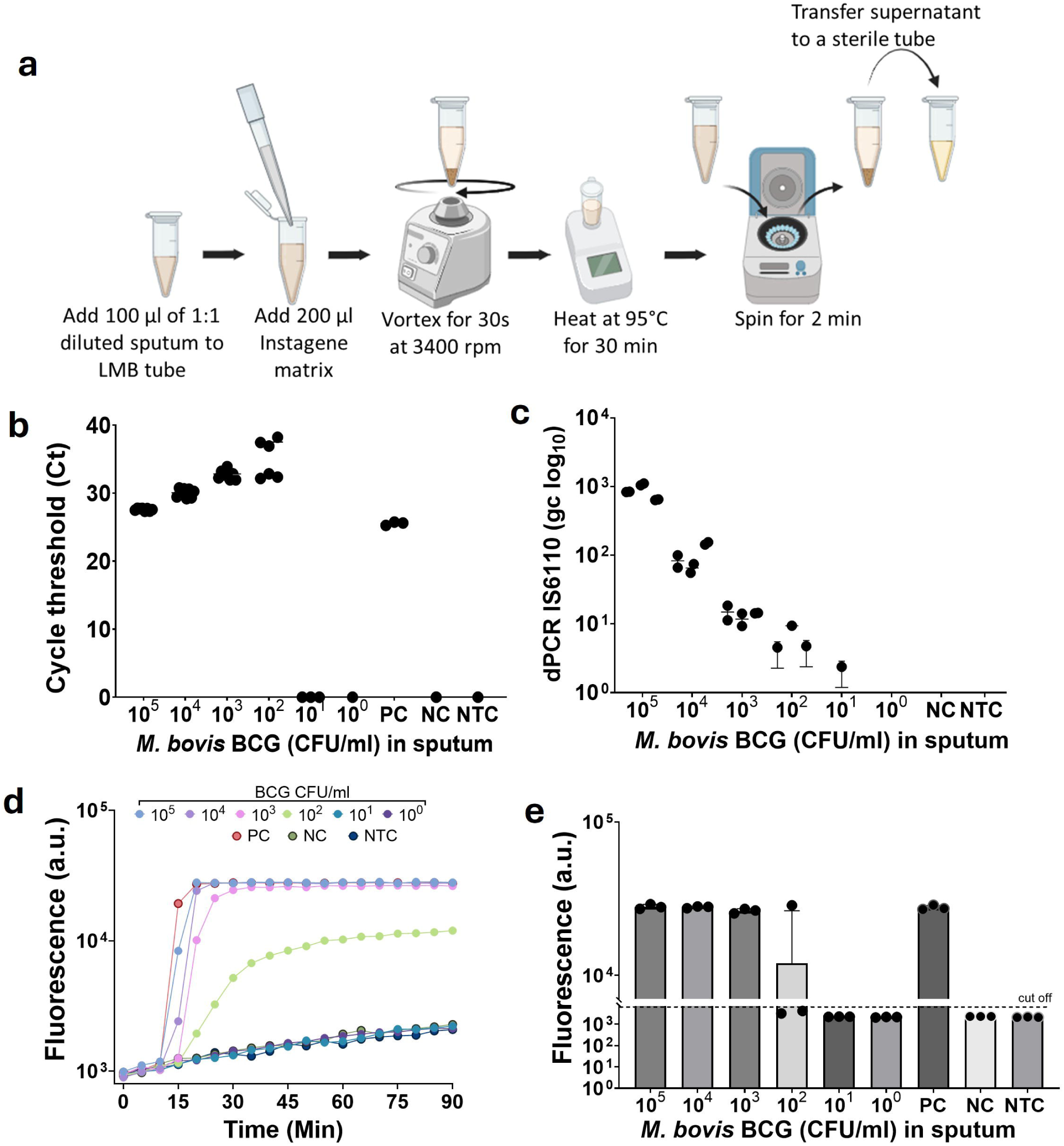
Sputum sample processing protocol evaluation with a single copy target *M. bovis* BCG IS6110. a) Final optimized sputum sample processing protocol; dynamic range for total DNA extracted from spiked pooled sputum was tested in b) real time PCR targeting IS6110 gene in Roche LC480 (N=8 with 3 biological replicates) c) Qiacuity dPCR targeting IS6110 (N=6 with 3 biological replicates and 2 technical replicates); c) and d) SHINE-TB cas13a assay in Cytation3 platform tested showing c) real time curves (FAM λEx. 490 nm/ λEm. 517nm). d) end point relative fluorescence units at 90 min (FAM λEx. 490 nm/ λEm. 517nm). PC-Positive control *M. bovis* BCG (at 2000 copies/reaction); NC-Extraction negative control; NTC-No template reaction control.

Extraction efficiency, calculated from the data using *M. bovis* BCG spiked into sputum at different concentrations, was found to be 48% at 10L CFU/ml, 62.2% at 10L CFU/ml, 86.4% at 10³ CFU/ml, and 99.3% at 10 CFU/ml, with an overall average extraction efficiency of approximately 74%.

### Co-extraction of *Mtb* and SARS-CoV-2 spiked in a single salivary sputum

*M. bovis* BCG and inactivated SARS-CoV-2 USA WA1/2020 (*SC2*) strains were spiked at 10^5^ CFU/ml or 10^5^ PFU/ml to pooled negative salivary sputum samples at different conditions. A set of samples (N=3) contained only *M. bovis* BCG and a second set contained only *SC2* and a third set contained both organisms spiked at 1:1 ratio. The samples were processed using the optimized sputum processing protocol as described in Fig. 4a.

All samples were evaluated using both dPCR and CRISPR SHINE-TB. As shown in Fig. 5, both BCG DNA (Fig. 5a, c) and *SC2* RNA (Fig. 5b, d), were efficiently extracted using our CRB protocol and detected using both dPCR and SHINE-TB.

**Fig 5.**
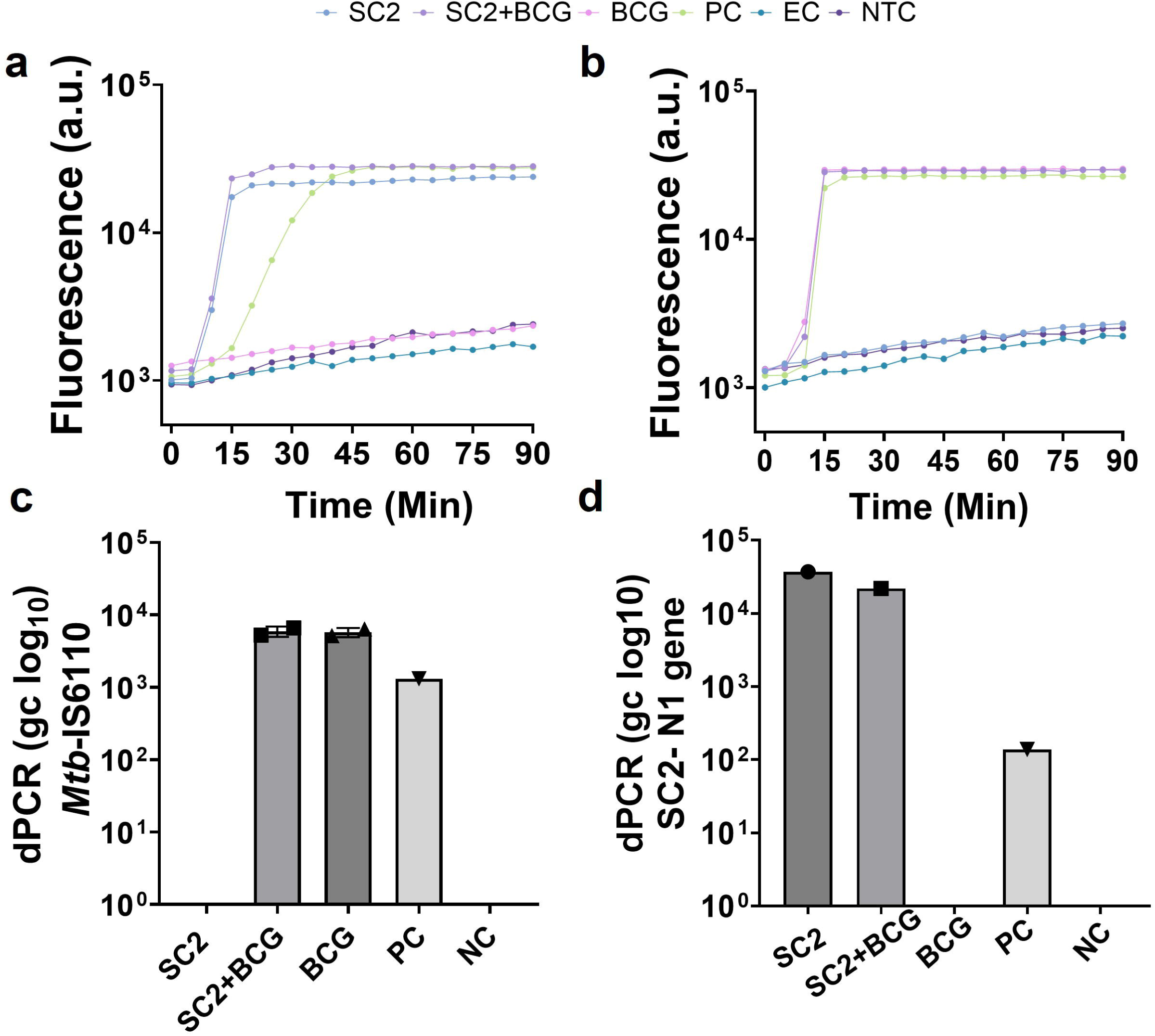
Co-detection of *Mtb* (10^5^ CFU/ml) and *SARS-CoV-2* (10^5^ PFU/ml) in salivary sputum (1:1 saliva to sputum) using CRISPR Cas13 assays for *Mtb* (a) and *SC2* (b) and dPCR for *Mtb* (c) and *SC2* (d). PC-Positive control *M. bovis* BCG (at 2000 copies/reaction) or *SARS-CoV-2* (*SC2*) WT strain WA1; NC-Extraction negative control; NTC-No template control.

## DISCUSSION

In this study, we have optimized and developed a simple, rapid and efficient sample processing method for total nucleic acid extraction from saliva and salivary sputum as a potential sample type for dual respiratory pathogen detection. This method utilizes Chelex-100 resin (Instagene Matrix) and a lysing matrix tube with 0.1 mm silica beads, combined with heating at 95°C, which demonstrated an extraction efficiency ranging from 48% (for 10^5^ CFU/ml) to as high as 99.3% (10 CFU/ml), which is better than the published methods (39, 40).

Considering *M. tuberculosis* (*Mtb*) is a hardy and difficult-to-lyse bacterium compared to *SC2*, our initial studies focused on optimizing *Mtb* lysis in both saliva and sputum. While sputum has been the primary specimen of choice for TB diagnosis, it presents various challenges for nucleic acid amplification tests. Alternative sample types such as saliva (18), oral swabs (41) and tongue swabs (42) have shown promise for respiratory pathogen detection. Saliva has been widely used for SARS-CoV-2 (*SC2*) diagnosis, which has shown significant variability in physical characteristics and potential contaminants (e.g., blood, drugs, food, tissue), necessitating careful optimization to maximize pathogen nucleic acid extraction efficiency. The choice of extraction protocol depends on the intended downstream application—while crude extracts suffice for most PCR-based diagnostics, higher-quality samples are required for whole-genome sequencing or transcriptomics.

For point-of-care (POC) diagnostic testing, we screened simple and low-cost methods that can be used by non-laboratory personnel. After a thorough literature search, we identified procedures and their limitations that overlap between standard viral and bacterial extraction methods and require minimal manipulation (Supplementary Table 1); ideally steps that are utilized in the field currently. Chelex-100 resin and heating seem to be commonly utilized for both *SC2* and *Mtb*, so these procedures were evaluated for our purposes. CRISPR-based diagnostic testing is emerging as a highly promising POC method for pathogen surveillance and strain identification (20, 43–45). However, comprehensive evaluation and optimization of sputum sample processing remain critical for TB detection.

We demonstrated that nucleic acids extracted using our method from *Mtb*-spiked saliva and sputum were compatible across different platforms, including LC480 qPCR, Qiacuity digital PCR (dPCR), and the CRISPR-based SHINE-TB test, detecting as low as 100 CFU/ml (Fig. 2, (38)). However, *Mtb*-spiked individual sputum samples showed lower compatibility, with an 82% positivity rate (18/22, Fig. 3d) compared to 100% detection using dPCR (Fig. 3c). The decreased detection rate in some samples (Sp-06, Sp-07, Sp-09, Sp-11) may be due to the presence of assay-specific inhibitors affecting the CRISPR test, such as recombinase polymerase amplification (RPA) or Cas enzymes, which require further investigation. Our preliminary tests with extracted negative matrices spiked with DNA did not show inhibition, and even the presence of up to 10% blood in saliva did not affect SHINE-TB performance (Fig. S2), suggesting effective removal of common inhibitors by our CRB protocol. To mitigate potential Cas/RPA enzyme inhibition, dilution with TE buffer (1:1) rescued 50% (2/4) of the inhibitory samples. Additionally, we explored mass-based filter tips (Monolith, C18 resin, C4 resin, strong cationic resin) to isolate genomic material from large macromolecules (e.g., lipids, proteins, food) (Fig. S3 A); PCR additives (Fig. S3 B); and varying concentrations of Chelex-100 resin (6%–20%, Fig. S3 C), but with limited success. Due to sample volume constraints, we could not repeat testing on all failed samples. These findings suggest that further sample cleanup may be necessary for some sputum samples to improve with CRISPR-based diagnostic tests. Further investigation is required to identify the inhibitors and develop targeted measures to mitigate their effects.

We also demonstrated our protocol’s ability to co-extract total nucleic acids (DNA and RNA) from *Mtb* and *SC2*, enabling dual detection using SHINE-TB (38) and SHINE-CoV-2 (37) assays (Fig. 5), as well as dPCR targeting the *N1* gene for *SC2* and *IS6110* for TB (Fig. 5). To our knowledge, this is the first reported co-extraction protocol from a single sample for simultaneous detection of both pathogens. Further work is underway to simplify the process, reduce processing time, improve sensitivity, and ensure nucleic acid stability. Additionally, we are currently investigating *Mtb* inactivation using the MGIT system.

In summary, we have successfully optimized a simple, rapid and efficient DNA/RNA co-extraction method for multi-pathogen testing, demonstrating compatibility across various diagnostic platforms. This co-extraction protocol for two very different pathogens may be adapted to other organisms to synergize TB and future pandemic surveillance and enhance routine multipathogen respiratory diagnostics. Ongoing optimization efforts will focus on expanding compatibility to additional respiratory specimens. The WHO is advocating for rapid and sensitive diagnostic approaches for pulmonary and extrapulmonary TB using non-sputum samples to enable same-day treatment initiation in resource-limited health clinics (46, 47). Future directions include eliminating or minimizing the need for specialized equipment, reducing time of incubation, enabling diagnostic testing directly in household settings for rapid medical diagnosis, and evaluating alternative sample types such as oral and tongue swabs.

## Data Availability

All data produced in the present study are presented here and as supplementary data. Any further information can be available upon reasonable request to the authors.

## ACKNOWLEDGEMENTS.

Funding was provided by the National Institutes of Health (NIH) R21 AI168808 (Y.L.X., P.B., C.M.), NIH R01 AI182281 (Y.L.X, C.M.), New Jersey Alliance for Clinical and Translational Science (NJACTS) UL1TR003017 (Y.L.X., P.B., C.M.) and Centers for Disease Control and Prevention 75D30122C15113 (C.M.). A.G.B. was supported by NIH Training Grants T32GM148739 and T32GM007388. We wish to thank Dr. David Alland for guidance and support.

## AUTHOR CONTRIBUTIONS

P.B., Y.L.X., C.M. conceptualized, supervised the study, and approved the final manuscript. N.M. performed the experiments and wrote the initial manuscript draft. P.B. finalized the manuscript with input from all authors. A.B, O.D., N.D., Y.H. performed supporting experiments and assisted with study implementation, A.W. and E.H. enrolled saliva and sputum donors.

## REFERENCES

1. Trajman A, Felker I, Alves LC, Coutinho I, Osman M, Meehan SA, Singh UB, Schwartz Y. 2022. The COVID-19 and TB syndemic: the way forward. Int J Tuberc Lung Dis 26:710–719.

2. WHO. 2024. Number of COVID-19 cases reported to WHO, Geneva.

3. WHO. 2024. Global tuberculosis report 2024. World Health Organization, Geneva.

4. Bagcchi S. 2023. WHO’s Global Tuberculosis Report 2022. Lancet Microbe 4:e20.

5. Pokam BDT, Shindoh EN, Djuikoue CI, Nana CS, Kakah SH, Elisee APK, Tendongfor N. 2024. The Effects of Coronavirus Disease-19 Pandemic on Tuberculosis Treatment Uptake and Outcomes in the Fako Division of Cameroon. Int J Mycobacteriol 13:387–393.

6. WHO. 2024. EpidemiologicalUpdate: SARS-CoV-2 and Other Respiratory Viruses in the Americas Region PAHO/WHO, Washington, D.C.

7. WHO. 2024. Tuberculosis resurges as top infectious disease killer.

8. Jeong Y, Min J. 2023. Impact of COVID-19 Pandemic on Tuberculosis Preventive Services and Their Post-Pandemic Recovery Strategies: A Rapid Review of Literature. J Korean Med Sci 38:e43.

9. Dheda K, Perumal T, Moultrie H, Perumal R, Esmail A, Scott AJ, Udwadia Z, Chang KC, Peter J, Pooran A, von Delft A, von Delft D, Martinson N, Loveday M, Charalambous S, Kachingwe E, Jassat W, Cohen C, Tempia S, Fennelly K, Pai M. 2022. The intersecting pandemics of tuberculosis and COVID-19: population-level and patient-level impact, clinical presentation, and corrective interventions. Lancet Respir Med 10:603–622.

10. Falzon D, Zignol M, Bastard M, Floyd K, Kasaeva T. 2023. The impact of the COVID-19 pandemic on the global tuberculosis epidemic. Front Immunol 14:1234785.

11. Ranasinghe L, Achar J, Gröschel MI, Whittaker E, Dodd PJ, Seddon JA. 2022. Global impact of COVID-19 on childhood tuberculosis: an analysis of notification data. Lancet Glob Health 10:e1774–e1781.

12. Genade LP, Kahamba T, Scott L, Tempia S, Walaza S, David A, Stevens W, Hlongwane K, von Gottberg A, Du Plessis M, Kleynhans J, Cohen C, Martinson NA. 2023. Co-testing a single sputum specimen for TB and SARS-CoV-2. Int J Tuberc Lung Dis 27:146–147.

13. Scott AJ, Limbada M, Perumal T, Jaumdally S, Kotze A, van der Merwe C, Cheeba M, Milimo D, Murphy K, van Ginneken B, de Kock M, Warren RM, Gina P, Swanepoel J, Kühn L, Oelofse S, Pooran A, Esmail A, Ayles H, Dheda K. 2024. Integrating molecular and radiological screening tools during community-based active case-finding for tuberculosis and COVID-19 in southern Africa. International Journal of Infectious Diseases 145:107081.

14. MacLean E, Kohli M, Weber SF, Suresh A, Schumacher SG, Denkinger CM, Pai M. 2020. Advances in Molecular Diagnosis of Tuberculosis. J Clin Microbiol 58.

15. Akowuah E, Acheampong G, Ayisi-Boateng NK, Amaniampong A, Agyapong FO, Senyo Kamasah J, Agyei G, Owusu DO, Nkrumah B, Mutocheluh M, Sylverken AA, Owusu M. 2022. Comparable Detection of SARS-CoV-2 in Sputum and Oropharyngeal Swab Samples of Suspected COVID-19 Patients. COVID 2:858–866.

16. Banada P, Elson D, Daivaa N, Park C, Desind S, Montalvan I, Kwiatkowski R, Chakravorty S, Alland D, Xie YL. 2021. Sample collection and transport strategies to enhance yield, accessibility, and biosafety of COVID-19 RT-PCR testing. J Med Microbiol 70.

17. Marín-Echeverri C, Pérez-Zapata L, Álvarez-Acevedo L, Gutiérrez-Hincapié S, Adams-Parra M, Tirado-Duarte D, Bolívar-Muñoz J, Gallego-Gómez M, Galeano-Castañeda Y, Piedrahita-Ochoa C, Del Valle Arrieta H. 2024. Diagnostic performance, stability, and acceptability of self-collected saliva without additives for SARS-CoV-2 molecular diagnosis. Eur J Clin Microbiol Infect Dis 43:1127–1138.

18. Ayalew S, Wegayehu T, Wondale B, Kebede D, Osman M, Niway S, Tarekegn A, Tessema B, Berg S, Ashford RT, Mihret A. 2024. Detection of *Mycobacterium tuberculosis* complex in saliva by quantitative PCR: A potential alternative specimen for pulmonary tuberculosis diagnosis. Tuberculosis 148:102554.

19. Namuganga AR, Chegou NN, Mubiri P, Walzl G, Mayanja-Kizza H. 2017. Suitability of saliva for Tuberculosis diagnosis: comparing with serum. BMC Infect Dis 17:600.

20. Arizti-Sanz J, Bradley AD, Zhang YB, Boehm CK, Freije CA, Grunberg ME, Kosoko-Thoroddsen T-SF, Welch NL, Pillai PP, Mantena S, Kim G, Uwanibe JN, John OG, Eromon PE, Kocher G, Gross R, Lee JS, Hensley LE, MacInnis BL, Johnson J, Springer M, Happi CT, Sabeti PC, Myhrvold C. 2022. Simplified Cas13-based assays for the fast identification of SARS-CoV-2 and its variants. Nature Biomedical Engineering 6:932–943.

21. Datta M, Singh DD, Naqvi AR. 2021. Molecular Diagnostic Tools for the Detection of SARS-CoV-2. Int Rev Immunol 40:143–156.

22. Kostyusheva A, Brezgin S, Babin Y, Vasilyeva I, Glebe D, Kostyushev D, Chulanov V. 2022. CRISPR-Cas systems for diagnosing infectious diseases. Methods 203:431–446.

23. Chakravorty S, Simmons AM, Rowneki M, Parmar H, Cao Y, Ryan J, Banada PP, Deshpande S, Shenai S, Gall A, Glass J, Krieswirth B, Schumacher SG, Nabeta P, Tukvadze N, Rodrigues C, Skrahina A, Tagliani E, Cirillo DM, Davidow A, Denkinger CM, Persing D, Kwiatkowski R, Jones M, Alland D. 2017. The New Xpert MTB/RIF Ultra: Improving Detection of *Mycobacterium tuberculosis* and Resistance to Rifampin in an Assay Suitable for Point-of-Care Testing. mBio 8.

24. Andrianto A, Mertaniasih NM, Gandi P, Al-Farabi MJ, Azmi Y, Jonatan M, Silahooij SI. 2020. Diagnostic test accuracy of Xpert MTB/RIF for tuberculous pericarditis: a systematic review and meta-analysis. F1000Res 9:761.

25. Promsena P, Jantarabenjakul W, Suntarattiwong P, Sudjaritruk T, Anugulruengkitt S, Rotcheewaphan S, Petsong S, Sawangsinth P, Sophonphan J, Tawan M, Moonwong J, Puthanakit T. 2022. Diagnostic Accuracy of Loop-Mediated Isothermal Amplification (TB-LAMP) for Tuberculosis in Children. J Pediatric Infect Dis Soc 11:9–15.

26. Sam IK, Chen YY, Ma J, Li SY, Ying RY, Li LX, Ji P, Wang SJ, Xu J, Bao YJ, Zhao GP, Zheng HJ, Wang J, Sha W, Wang Y. 2021. TB-QUICK: CRISPR-Cas12b-assisted rapid and sensitive detection of *Mycobacterium tuberculosis*. J Infect 83:54–60.

27. Yan L, Xiao H, Zhang Q. 2016. Systematic review: Comparison of Xpert MTB/RIF, LAMP and SAT methods for the diagnosis of pulmonary tuberculosis. Tuberculosis (Edinb) 96:75–86.

28. Loeffelholz MJ, Alland D, Butler-Wu SM, Pandey U, Perno CF, Nava A, Carroll KC, Mostafa H, Davies E, McEwan A, Rakeman JL, Fowler RC, Pawlotsky JM, Fourati S, Banik S, Banada PP, Swaminathan S, Chakravorty S, Kwiatkowski RW, Chu VC, Kop J, Gaur R, Sin MLY, Nguyen D, Singh S, Zhang N, Persing DH. 2020. Multicenter Evaluation of the Cepheid Xpert Xpress SARS-CoV-2 Test. J Clin Microbiol 58.

29. Banada PP, Green R, Streck D, Kurvathi R, Reiss R, Banik S, Daivaa N, Montalvan I, Jones R, Marras SAE, Chakravorty S, Alland D. 2023. An expanded RT-PCR melting temperature coding assay to rapidly identify all known SARS-CoV-2 variants and sub-variants of concern. Sci Rep 13:21927.

30. Banada P, Green R, Banik S, Chopoorian A, Streck D, Jones R, Chakravorty S, Alland D. 2021. A Simple RT-PCR Melting temperature Assay to Rapidly Screen for Widely Circulating SARS-CoV-2 Variants. medRxiv doi:10.1101/2021.03.05.21252709.

31. Broger T, Marx FM, Theron G, Marais BJ, Nicol MP, Kerkhoff AD, Nathavitharana R, Huerga H, Gupta-Wright A, Kohli M, Nichols BE, Muyoyeta M, Meintjes G, Ruhwald M, Peeling RW, Pai NP, Pollock NR, Pai M, Cattamanchi A, Dowdy DW, Dewan P, Denkinger CM. 2024. Diagnostic yield as an important metric for the evaluation of novel tuberculosis tests: rationale and guidance for future research. Lancet Glob Health 12:e1184–e1191.

32. Aldous WK, Pounder JI, Cloud JL, Woods GL. 2005. Comparison of six methods of extracting *Mycobacterium tuberculosis* DNA from processed sputum for testing by quantitative real-time PCR. J Clin Microbiol 43:2471–3.

33. Chakravorty S, Tyagi JS. 2001. Novel use of guanidinium isothiocyanate in the isolation of *Mycobacterium tuberculosis* DNA from clinical material. FEMS Microbiol Lett 205:113–7.

34. Koentjoro MP, Donastin A, Prasetyo EN. 2021. A simple method of dna extraction of *Mycobacterium tuberculosis* from sputum cultures for sequencing analysis. Afr J Infect Dis 15:19–22.

35. Thakore N, Norville R, Franke M, Calderon R, Lecca L, Villanueva M, Murray MB, Cooney CG, Chandler DP, Holmberg RC. 2018. Automated TruTip nucleic acid extraction and purification from raw sputum. PLoS One 13:e0199869.

36. Malherbe ST, Shenai S, Ronacher K, Loxton AG, Dolganov G, Kriel M, Van T, Chen RY, Warwick J, Via LE, Song T, Lee M, Schoolnik G, Tromp G, Alland D, Barry CE, 3rd, Winter J, Walzl G, Lucas L, Spuy GV, Stanley K, Thiart L, Smith B, Du Plessis N, Beltran CG, Maasdorp E, Ellmann A, Choi H, Joh J, Dodd LE, Allwood B, Koegelenberg C, Vorster M, Griffith-Richards S. 2016. Persisting positron emission tomography lesion activity and *Mycobacterium tuberculosis* mRNA after tuberculosis cure. Nat Med 22:1094–1100.

37. Lu X, Wang L, Sakthivel SK, Whitaker B, Murray J, Kamili S, Lynch B, Malapati L, Burke SA, Harcourt J, Tamin A, Thornburg NJ, Villanueva JM, Lindstrom S. 2020. US CDC Real-Time Reverse Transcription PCR Panel for Detection of Severe Acute Respiratory Syndrome Coronavirus 2. Emerg Infect Dis 26:1654–65.

38. Dunkley ORS, Bell AG, Modi NH, Huang Y, Tseng S, Reiss R, Daivaa N, Davis JL, Vargas DA, Banada P, Xie YL, Myhrvold C. 2025. A Streamlined Point-of-Care CRISPR Test for Tuberculosis Detection Directly from Sputum. medRxiv doi:10.1101/2025.02.19.25322517:2025.02.19.25322517.

39. Kolia-Diafouka P, Godreuil S, Bourdin A, Carrère-Kremer S, Kremer L, Van de Perre P, Tuaillon E. 2018. Optimized Lysis-Extraction Method Combined With IS6110-Amplification for Detection of *Mycobacterium tuberculosis* in Paucibacillary Sputum Specimens. Frontiers in Microbiology 9.

40. Pan S, Gu B, Wang H, Yan Z, Wang P, Pei H, Xie W, Chen D, Liu G. 2013. Comparison of four DNA extraction methods for detecting *Mycobacterium tuberculosis* by real-time PCR and its clinical application in pulmonary tuberculosis. Journal of Thoracic Disease 5:251–257.

41. Church EC, Steingart KR, Cangelosi GA, Ruhwald M, Kohli M, Shapiro AE. 2024. Oral swabs with a rapid molecular diagnostic test for pulmonary tuberculosis in adults and children: a systematic review. Lancet Glob Health 12:e45–e54.

42. Steadman A, Andama A, Ball A, Mukwatamundu J, Khimani K, Mochizuki T, Asege L, Bukirwa A, Kato JB, Katumba D, Kisakye E, Mangeni W, Mwebe S, Nakaye M, Nassuna I, Nyawere J, Nakaweesa A, Cook C, Phillips P, Nalugwa T, Bachman CM, Semitala FC, Weigl BH, Connelly J, Worodria W, Cattamanchi A. 2024. New Manual Quantitative Polymerase Chain Reaction Assay Validated on Tongue Swabs Collected and Processed in Uganda Shows Sensitivity That Rivals Sputum-based Molecular Tuberculosis Diagnostics. Clinical Infectious Diseases 78:1313–1320.

43. Arizti-Sanz J, Freije CA, Stanton AC, Petros BA, Boehm CK, Siddiqui S, Shaw BM, Adams G, Kosoko-Thoroddsen T-SF, Kemball ME, Uwanibe JN, Ajogbasile FV, Eromon PE, Gross R, Wronka L, Caviness K, Hensley LE, Bergman NH, MacInnis BL, Happi CT, Lemieux JE, Sabeti PC, Myhrvold C. 2020. Streamlined inactivation, amplification, and Cas13-based detection of SARS-CoV-2. Nature Communications 11:5921.

44. de Puig H, Lee RA, Najjar D, Tan X, Soenksen LR, Angenent-Mari NM, Donghia NM, Weckman NE, Ory A, Ng CF, Nguyen PQ, Mao AS, Ferrante TC, Lansberry G, Sallum H, Niemi J, Collins JJ. 2021. Minimally instrumented SHERLOCK (miSHERLOCK) for CRISPR-based point-of-care diagnosis of SARS-CoV-2 and emerging variants. Science Advances 7:eabh2944.

45. Joung J, Ladha A, Saito M, Kim N-G, Woolley AE, Segel M, Barretto RPJ, Ranu A, Macrae RK, Faure G, Ioannidi EI, Krajeski RN, Bruneau R, Huang M-LW, Yu XG, Li JZ, Walker BD, Hung DT, Greninger AL, Jerome KR, Gootenberg JS, Abudayyeh OO, Zhang F. 2020. Detection of SARS-CoV-2 with SHERLOCK One-Pot Testing. New England Journal of Medicine 383:1492–1494.

46. Kasule GW, Hermans S, Semugenze D, Wekiya E, Nsubuga J, Mwachan P, Kabugo J, Joloba M, García-Basteiro AL, Ssengooba W, Elisa L-V, Belén S-C, Lucía C-C, Sergi S, Ehrlich J, Fernandez C, Makhosazana D, Gcinile D, Nomathemba D, Nkulungwane M, Nokwanda K, Mbongeni D, Busizwe S, Ziyane M, Mulengwa D, Adu-Gyamfi CG, Maphalala N, Babongile N, Shiba N, Dlamini F, Shabalala F, Dlamini S, Maphalala G, Dlamini L, Dube S, Acacio S, Munguambe S, Fonseca LJ, Cumbe M, Mambuque ET, Lima A, Magul K, Tembe G, Mudumane BV, Cebola F, Rungo J, Junior AB, Gomis N, Nassolo M, Wobudeya E, et al. 2024. Non-sputum-based samples and biomarkers for detection of *Mycobacterium tuberculosis*: the hope to improve childhood and HIV-associated tuberculosis diagnosis. European Journal of Medical Research 29:502.

47. WHO. 2024. Target product profiles for tuberculosis diagnosis and detection of drug resistance. World Health Organization,

