## Supplementary materials for "Simplified Co-extraction of total Nucleic Acids from Respiratory Samples for detection of *Mycobacterium tuberculosis* and SARS-CoV-2 optimized for compatibility across Diagnostic Platforms"

**Supplementary Table 1**. Literature Search of common methodologies and limitations of existing procedures for *Mtb* and *SC2* nucleic acid extraction.

| **No.** | **Reference** | **Extraction method** | **Notes** | **Processing time** | **Limitations or different from our method** |
| --- | --- | --- | --- | --- | --- |
| **For DNA extraction from *Mycobacterium tuberculosis (Mtb)*** | | | | | |
| 1 | (34) | Boil method | In this simple method, distilled water was added to the sputum sample, mixed via vortex and placed in boiling water bath (100°C) for 3 min. The mixture was then centrifuged at 15000 rpm for 5 min at 4°C, and the supernatant was separated for DNA analysis using 1% agarose gel electrophoresis, PCR targeting *ripA* gene of *Mtb*, and sequencing. | ~8 min | Used for cultures not directly from samples  High speed centrifugation is needed. |
| 2 | (32) | TE Boil extraction | This method involves addition of TE buffer and placing the sample pellet and TE mixture in boiling water bath for 15 min followed by centrifugation at 16000xg for 5 min. The eluant was compatible with PCR. | ~20 min | Used for sample pellets, not directly from the samples requiring additional processing.  Need for specialized instruments such as vortex genie  Qiagen kits are expensive, and the procedures are long. Requires some technical skill. |
| 3 |  | IDI extraction | TE buffer and pellet was placed into an IDI lysis tube which contains glass bead matrix. The tube was vigorously mixed for 5 min using Vortex Genie 2 and then placed in a boiling water bath . | ~25 min |  |
| 4 |  | Qiagen extraction | In this method, authors used QIAGEN QIAmp DNA mini kit tissue protocol but added enzymatic digestion steps with lysozyme. Boiled for 15 min, and proteinase K step was at 56°C for 1 hour. Mean Ct values with the real time assays were like TE boil extraction method, however yield was lower. | >1h |  |
| 7 | (33) | GITC-boil prep method | In this method, diverse samples were preprocessed first in several different ways, involving several reagents such as *N*-acetyl L-cysteine, guanidinium isothiocyanate, Tris-HCl, EDTA, Sarcosyl, and M β-mercaptoethanol, and centrifugation, and 1-mm glass beads. After inhibitor removal step,Chelex-100 resin, Tween20 and Triton X-100 were added to the preprocessed samples. The mixture was then heated at 90°C for 40 min, followed by centrifugation. This method was tested with various pulmonary and extrapulmonary samples. DNA samples were compatible with PCR. | 3h | Very lengthy protocol,  Guanidium iso thiocyanate (GITC) is hazardous, produces toxic gas, difficult to discard easily.  Not compatible with point of care use with limited technical skills.  GITC samples may not be compatible with many non-PCR based downstream applications |
| 8 | (48) | Chelex-100 method combined with boiling and sonication | Sample culture pellet was incubated with 20%Chelex-100 resin. After vortex mixing, the mixture was placed for boiling at 100°C for 15 min, and placed in ultrasonic water bath for 15 min. DNA was separated after 5 min centrifugation at 14000g and was analyzed based on *IS6110* gene by real time qPCR in the LightCycler 480 system for efficiency. The proportion of *Mtb* recovered was 82% with this method relative to the estimated input, and authors chose to evaluate the clinical sputum samples with this method. | ~30 min | Culture pellet only. No direct sample.  Extraction efficiency was similar to our method.  Compatibility shown only for PCR based assays.  GITC is hazardous and not compatible with non-PCR based methods such as CRISPR based. |
| 9 |  | Boiling method combined with Guanidinium Isothiocyanate | Incubate sample pellet with lysis buffer containing Tris-HCl, EDTA, NaCl, and guanidium isothiocyanate (GTIC) for 20 min, and apply it through 3 cycles of freeze thawing (-80°C for 5 min and 100°C for 5 min) and boiling at 100°C for 15 min. DNA was analyzed for *IS6110* gene in PCR. The proportion of *Mtb* recovered was ~45% with this method relative to the estimated input. | >1h |  |
| 10 | (49) | MagPurix TB DNA extraction kit | Samples were processing to get pellets using NaLC-NaoH method. ComparedChelex-100 instagene matrix with MagPurix, a magnetic paticle separation technology and Magpurix 12s automated nucleic acid purification system using manufacturer’s recommendations. | 1h | Need for specialized kits and instruments.  Requires some technical skills. |
| 11 | (50) | Mechanical disruption with beads followed by boiling andChelex extraction | Different types of beads such as 0.1mm zirconia, 0.2mm and 1mm glass beads were mixed with sputum sample pellet suspended in TE. The mixture was vortexed for 5 min and incubated at 95°C for 5 min. 100µL debris free lysate after centrifugation was mixed with equal volume 10%Chelex 100 resins, incubated at room temperature for 10 min, and centrifuged at 1000g for 2 min. DNA was analyzed by multiplex PCR targeting *IS6110* and *Rv0927c-pstS3* regions of *Mtb* in Applied Biosystems 7700 real-time system. It was shown that smaller size glass beads result in better extraction with this method. | ~22 min | Pellets of samples. needs additional pre-processing. |
| **SARS-CoV-2 RNA extraction** | | | | | |
| 1 | (51) | Extraction by heat fixation and or using ProteinaseK | Saliva and oral swab samples were added with equal volume TE buffer or with Proteinase K, was incubated at room temperature for 10 min, followed by incubation at 98°C for 30-45 min. Extracted RNAs were evaluated by RT-PCR test targeting E and RdRp genes. For saliva, this method shows 71.43% diagnostic sensitivity, vs 56.12% for dry swab. | ~55 min | Lengthy, RT-PCR testing only.  No other downstream applications tested. |
| 2 | (52) | Acid pH-based method | This extraction method uses lysis buffer with pH 5 and containing SDS, Sodium citrate dehydrate, anhydrous citric acid, and EDTA. SDS lyse viral protein coats, and low pH helps in the recovery of RNA. After lysis buffer was added to the sample and mixed, precipitation buffer that also contains Sodium citrate dehydrate, anhydrous citric acid, and NaCl was added. The following steps are incubation on ice, multiple centrifugations, and supernatant recovery in isopropanol and 70% ethanol, and pellet resuspension in pre-warmed nuclease free water. Nasopharyngeal samples underwent this extraction, and the extracted RNA was evaluated by one step RT-qPCR targeting RNAseP, N1 and N2 genes. This method gave high yield and comparable results with commercial extraction kit. | ~40 min | NP specimens only  Not so simple.  SDS containing samples are not always compatible with downstream applications.  Requires many chemicals and reagents and multiple centrifugation steps, making this impractical for use in resource limited settings. |
| 3 | (53) | Guanidinium Isothiocyanate, phenol, and chloroform-based extraction | This method involves mixing the sample with three reagents guanidinium isothiocyanate, phenol, and chloroform, vortexing, incubation at room temperature, and centrifugation for 15 min at 12000g. RNAs were extracted from nasopharyngeal (NP) and oropharyngeal (OP) swab samples and evaluated by one-step RT-qPCR targeting N gene of SARS-CoV2. The detection level with this method was ~4copies/µL of RNA. | ~19 min | GITC based methods are not always compatible with non-PCR based diagnostics.  Introduces additional chemical hazard and difficult to easily discard samples due to potential toxic gases produced with common disinfectants like bleach. |

**Supplementary Table 2.** Primers and probes used.

| **Reference** | **Target gene (Organism)** | **Primer** | **5'** | **Sequence** | **3'** |
| --- | --- | --- | --- | --- | --- |
| Chakravorty et al., (23) | *IS6110 (Mtb)* | Forward |  | CGCCGCTTCGGACCACCAGCAC |  |
|  |  | Reverse |  | GTGACAAAGGCCACGTAGGCGAACC |  |
|  |  | Probe | FAM | CGGCTGTGGGTAGCAGACCTCACC | BHQ1 |
| Malherbe et al., (36) | *acr (Mtb)* | Forward |  | CTAATACCGGATAGGACCACGG |  |
|  |  | Reverse |  | CTCATCCCACACCGCTAAAGCG |  |
|  |  | Probe | FAM | CGCTCCCCTTCGTTCGCACGGTGTCGCTG GAGCG | DABCYL |
| Lu et al., (37) | *N1 (SARS-CoV-2)* | Forward |  | GACCCCAAAATCAGCGAAAT |  |
|  |  | Reverse |  | TCTGGTTACTGCCAGTTGAATCTG |  |
|  |  | Probe | FAM | ACCCCGCATTACGTTTGGTGGACC | BHQ1 |

**Supplementary Table 3. Participant Profile and Sputum Characteristics.** Characteristics of sputum (and saliva) donors and their sputum samples. Participants age ranged from 27 to 84 years.

T= thin, V=viscous, VV = very viscous, SV=slightly viscous, NA= not assessed

| **SpID** | **Race** | **Sex** | **Diagnosis** | **Sputum vol. (mL)** | **Sputum consistency** | **Sputum blood** | **Sputum color** |
| --- | --- | --- | --- | --- | --- | --- | --- |
| SP-01  SP-13 | Black | M | Pneumonia | 3.8  5 | V  V | no  no | clear  clear |
| SP-02 | Black | F | COPD | 3 | V | yes | tan |
| SP-03 | Black | F | Asthma | 7 | VV | no | clear |
| SP-04 | Black | M | Pyelonephritis | 6 | V | no | clear |
| SP-05 | Black | F | Interstitial lung disease | 4 | VV | no | clear |
| SP-06  SP-07 | Hispanic or Latino | F | Asthma | 5.5 | V  SV | no  no | white  tan |
| SP-08 | Black | M | Foot laceration | 5 | VV | no | other |
| SP-09  SP-10 | Black | F | Pneumonia, Asthma | 48  7 | VV  VV | yes  no | brown/red  tan |
| SP-11 | Black | M | Congestive heart failure | 3.5 | NA | NA | NA |
| SP-12 | Black | M | Congestive heart failure | 3.5 | NA | NA | NA |
| SP-14 | Black | F | COPD | 3 | VV | no | tan |
| SP-15 | Black | F | Asthma | 1 | VV | no | other |
| SP-16 | White | M | Femoral neck fracture | 3 | VV | no | tan |
| SP-17 | Black | M | COPD | 3 | VV | no | pink |
| SP-18 | Black | F | COPD, asthma | 3 | SV | no | white |
| SP-19 | Hispanic or Latino | M | COPD | 3 | VV | no | tan |
| SP-20 | Hispanic or Latino | M | Lung Kaposi Sarcoma | 3.4 | VV | yes | clear |
| SP-21 | Hispanic or Latino | F | Granulomatosis with polyangiitis | 3 | SV | no | tan |
| SP-22 | Hispanic or Latino | M | Mycobacterium avium complex pneumonia | 4.1 | V | yes | tan |

Fig. S1. **Instagene matrix and lab-madeChelex at different concentration perform equally for salivary sputum.** Different concentration ofChelex resin ranging from, 6% to 20% compared to Instagene matrix (IGM, Biorad) and in presence or absence of lysing matrix beads (LMBs) was explored for sputum diluted 1:1.

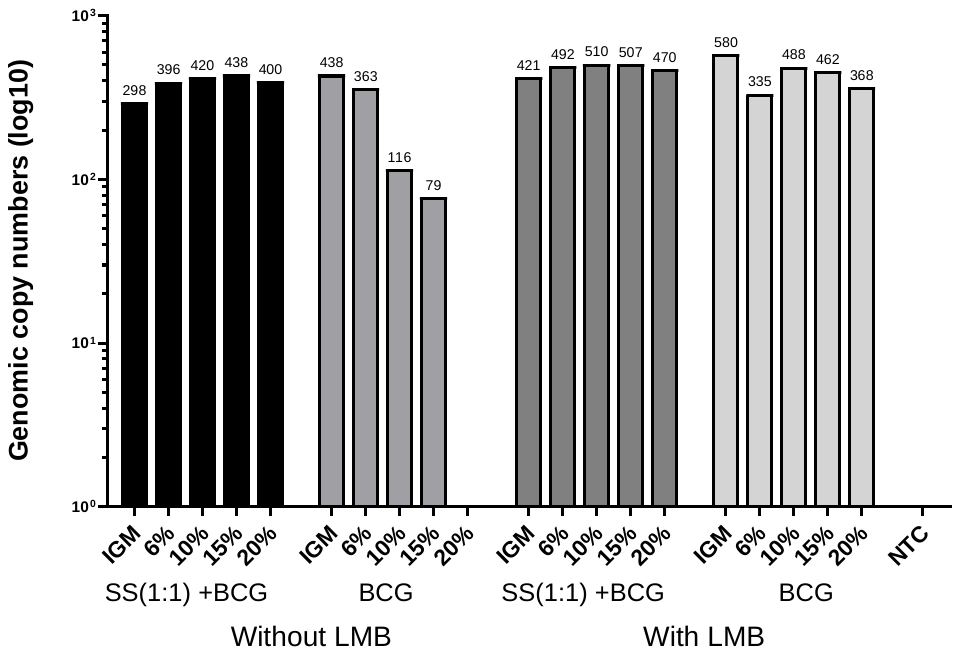

**A**

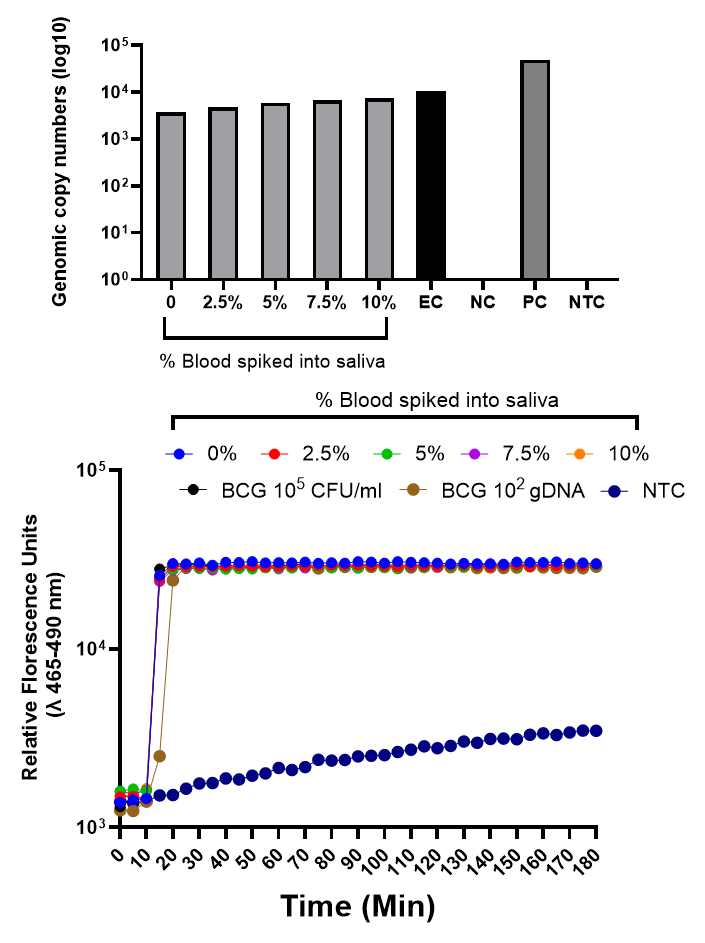

**Fig. S2. Efficient removal of inhibitors using CRB protocol.** *M. bovis* BCG spiked in saliva at 10^5^ CFU/ml was titrated with K2 EDTA blood spiked at different concentrations ranging from 0,2.5, 5, 7.5 and 10%. Total nucleic acids were extracted using the CRB protocol and the eluate was tested in both A) dPCR and B) CRISPR SHINE-TB test.

**B**

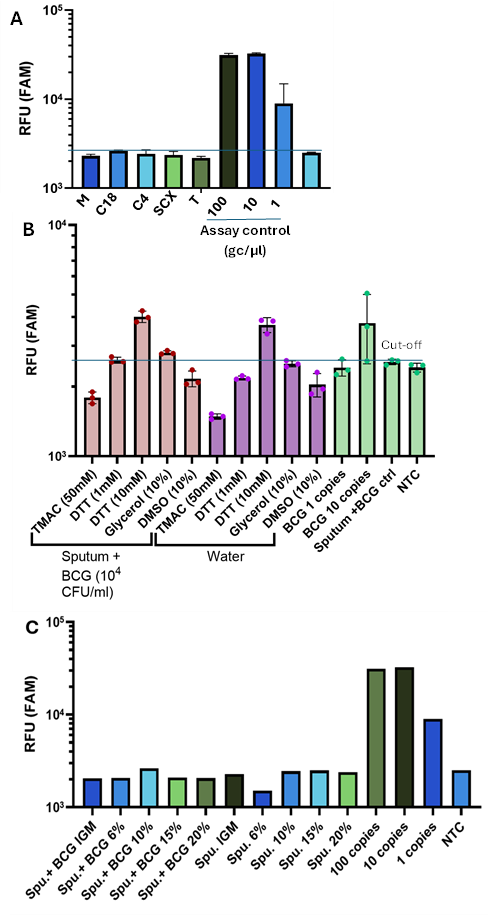

**Fig. S3. Removal of sputum inhibitors using different methods.** M. bovis BCG spiked in inhibitory pooled sputum at 10^5^ CFU/ml, extracted using CRB method and treated with A) mass-based carbon reverse phase micropipette tips; B) PCR additives and C) increased concentrations of Chelex-100 reagent; and tested in CRISPR SHINE-TB assay. Cut-off was established based on the NTC+ SD. M=Monolith, Cole-Parmer SPE; C18= carbon 18 filter tips, Pierce; C4= Ziptip, Millipore Sigma; SCX= Strong cation resin, Merck ZipTip; IGM= Biorad Instagene matrix; spu=sputum NTC=No template control. gc/µl= genomic copies per microliter.
